# No association between the SARS-CoV-2 variants and mortality rates in the Eastern Mediterranean Region

**DOI:** 10.1101/2021.01.06.21249332

**Authors:** Saad Omais, Samer Kharroubi, Hassan Zaraket

## Abstract

As the novel coronavirus SARS-CoV-2 continues to spread in all countries, there is a growing interest in monitoring and understanding the impact of emerging strains on virus transmission and disease severity. Here, we analyzed SARS-CoV-2 genomic sequences reported in the Eastern Mediterranean Region (EMR) countries, as of 1 January 2021. The majority (∼75%) of these sequences originated from three out of 22 EMR countries, and 65.8% of all sequences belonged to GISAID clades GR, GH, G and GV. A delay ranging between 30-150 days from sample collection to sequence submission was observed across all countries, limiting the utility of such data in informing public health policies. We identified ten common non-synonymous mutations represented among SARS-CoV-2 in the EMR and several country-specific ones. Two substitutions, spike_D614G and NSP12_P323L, were predominantly concurrent in most countries. While the single incidence of NSP12_P323L was positively correlated with higher case fatality rates in EMR, no such association was established for the double (spike_D614G and NSP12_P323L) concurrent variant across the region. Our study identified critical data gaps in EMR highlighting the importance of enhancing surveillance and sequencing capacities in the region.

## Introduction

In early October 2020, the World Health Organization (WHO) estimated that 10% of the global population has been infected with the novel coronavirus, SARS-CoV-2 (Keaten, 2020). Since its declaration as a pandemic seven months earlier, COVID-19 has inflicted substantial health and economic burden around the world (Fineberg, 2020; Ioannidis, 2020a). As of 22 January 2021, there have been over 96 million confirmed cases and 2 million deaths attributed to COVID-19 (WHO). Specifically, 5,461,398 confirmed cases (of whom 4,815,552 have recovered) and 130,079 deaths were reported in the Eastern Mediterranean Region (EMR) (WHO).

Prior to SARS-CoV-2 emergence, six human coronavirus (HCoV) strains were known to infect humans: two of which caused fatal respiratory diseases (SARS-CoV-1 and MERS-CoV), while the remaining four circulate annually and cause 15-29% of all common colds (HCoV-229E, - OC43, -NL63 and -HKU1) (Su et al., 2016). SARS-CoV-2 gains access to the host cell by binding to the human angiotensin-converting enzyme 2 (hACE2) through its spike (S) protein. SARS-CoV-2 S receptor-binding domain (RBD) has a 10-to 20-fold higher hACE2 binding affinity than SARS-CoV-1 (Wrapp et al., 2020). Yet, SARS-CoV-2 RBD is less accessible such that the overall S-hACE2 interaction is similar or weaker than that of SARS-CoV-1. Nonetheless, the SARS-CoV-2 spike protein possesses a polybasic cleavage site making it accessible to furin protease which enhances cell entry (Shang et al., 2020). Similar to other beta-coronaviruses, SARS-CoV-2 has a positive-sense RNA genome that is around 30-kb in length with six functional open reading frames (ORFs). ORFa and -b constitute nearly two-thirds of the genome and produce 16 non-structural proteins (NSPs), including the RNA-dependent RNA polymerase (RdRp or NSP12) and a helicase (NSP13). The structural nucleocapsid (N), membrane (M), envelope (E), and S proteins are encoded by the remaining stretch of the RNA genome along with other accessory proteins (Hartenian et al., 2020).

There has been a tremendous global surveillance effort to closely monitor SARS-CoV-2 circulating worldwide that enabled timely detection of emerging variants. SARS-CoV-2 evolves at an estimated rate of around 6 × 10^−4^ nucleotides/genome/year (van Dorp et al., 2020a). Numerous variants have emerged but only a handful of them have been fixated such as the S_D614G that has prevailed globally. S_D614G was shown to alter infectivity and virulence of SARS-CoV-2 and was associated with an increase in mortality (Eaaswarkhanth et al., 2020; Plante et al., 2020; van Dorp et al., 2020a; Vankadari, 2020; Wang et al., 2020). Specifically, Sallam et al. reported an increase in the incidence of S_D614G variant between February and June 2020 in the Middle East and North Africa (Sallam et al., 2021). However, a detailed genomic characterization of SARS-CoV-2 in EMR is still lacking. It also remains not clear whether the case fatality rates (CFRs) in EMR are associated with any of the prevailing SARS-CoV-2 variants. In this study, we analyzed SARS-CoV-2 whole-genome sequences collected in EMR and examined the temporal and country-level associations between the predominant variants and CFRs.

## Methods

### Sequence Retrieval and Analysis

SARS-CoV-2 sequences were downloaded from GISAID EpiCoV™ database (Shu and McCauley, 2017) which were submitted by 1 January 2021 from EMR countries. These are Afghanistan, Bahrain, Djibouti, Egypt, Iran, Iraq, Jordan, Kuwait, Lebanon, Libya, Morocco, Oman, Pakistan, Palestine, Qatar, Saudi Arabia, Somalia, Sudan, Syrian Arab Republic, Tunisia,

United Arab Emirates (UAE), and Yemen (http://www.emro.who.int/countries.html). Only ‘high coverage’ sequences were selected, i.e. “entries with <1% Ns and <0.05% unique amino acid mutations (not seen in other sequences in the database) and no insertion/deletion unless verified by submitter”. These were then analyzed by CoVsurver enabled by GISAID (https://www.gisaid.org/epiflu-applications/covsurver-mutations-app/) and compared with reference strain hCoV-19/Wuhan/WIV04/2019 (accession number: EPI_ISL_402124). Sequence metadata corresponding to age and gender were extracted from EpiCoV™ ‘nextmeta’ file for sequences submitted as of 12 November 2020. For clade analysis, we used the Nextclade tool from the Nextstrain project (https://clades.nextstrain.org/) (Hadfield et al., 2018) and GISAID clade designation provided by CoVsurver analysis.

### Country Data and Mortality Rate

Data on population size, total COVID-19 cases, and total deaths were retrieved from the Worldometers website as of 22 January 2021 (https://www.worldometers.info/coronavirus/). Data on daily cases and deaths per country were extracted from WHO Coronavirus Disease (COVID-19) Dashboard (https://covid19.who.int/) (WHO). CFR was calculated and adjusted for a 13-day lag time from reporting to death, such that the denominator was designated as the cumulative cases from 13 days earlier, while the numerator totaled the cumulative deaths till 13 days earlier plus half the additional deaths recorded during the lag period (Wilson et al., 2020).

### Statistical Analysis

The data were checked for completeness and entered into the Statistical Package for the Social Sciences (SPSS) software version 20 for Windows, which was later used for statistical analyses [IBM: Statistical Package for the Social Sciences. SPSS Statistics 2013, 20.]. Eight of the 15 EMR countries that have sequence data available were excluded from cross-country fatality correlations for the following reasons: Egypt had a relatively high CFR (5.9%) probably as a result of the low number of reported cases (1073 cases per million) likely due to limited testing capacity; Iraq, Pakistan, Tunisia, Lebanon, Kuwait, and Qatar had a relatively low sequence count each (<50, 1.5% of all sequences) (**Figure 1A**), Iran had only 18 complete sequences (>25 kbp) out of 104 sequences and was thus less comparable to other sequences (**Supplementary Figure 1)**. For the summary of the data, descriptive statistics were presented to summarize the study variables of interest as counts and percentages across countries. Pie charts and bar plots were used to chart comparison in count and percentages of variants across all countries. An independent t-test was used to chart the comparison of variants via gender (male vs female), while one-way ANOVA was carried out for age distribution (0-9, 10-18, 19-49, 50-59, and >60). Pearson’s correlation coefficients were used to examine the association between variant percentages and CFR for the seven countries considered in this study (Toyoshima et al., 2020). For all analyses done, a *p-value* of less than 0.05 was used to detect statistical significance.

**Figure 1.**
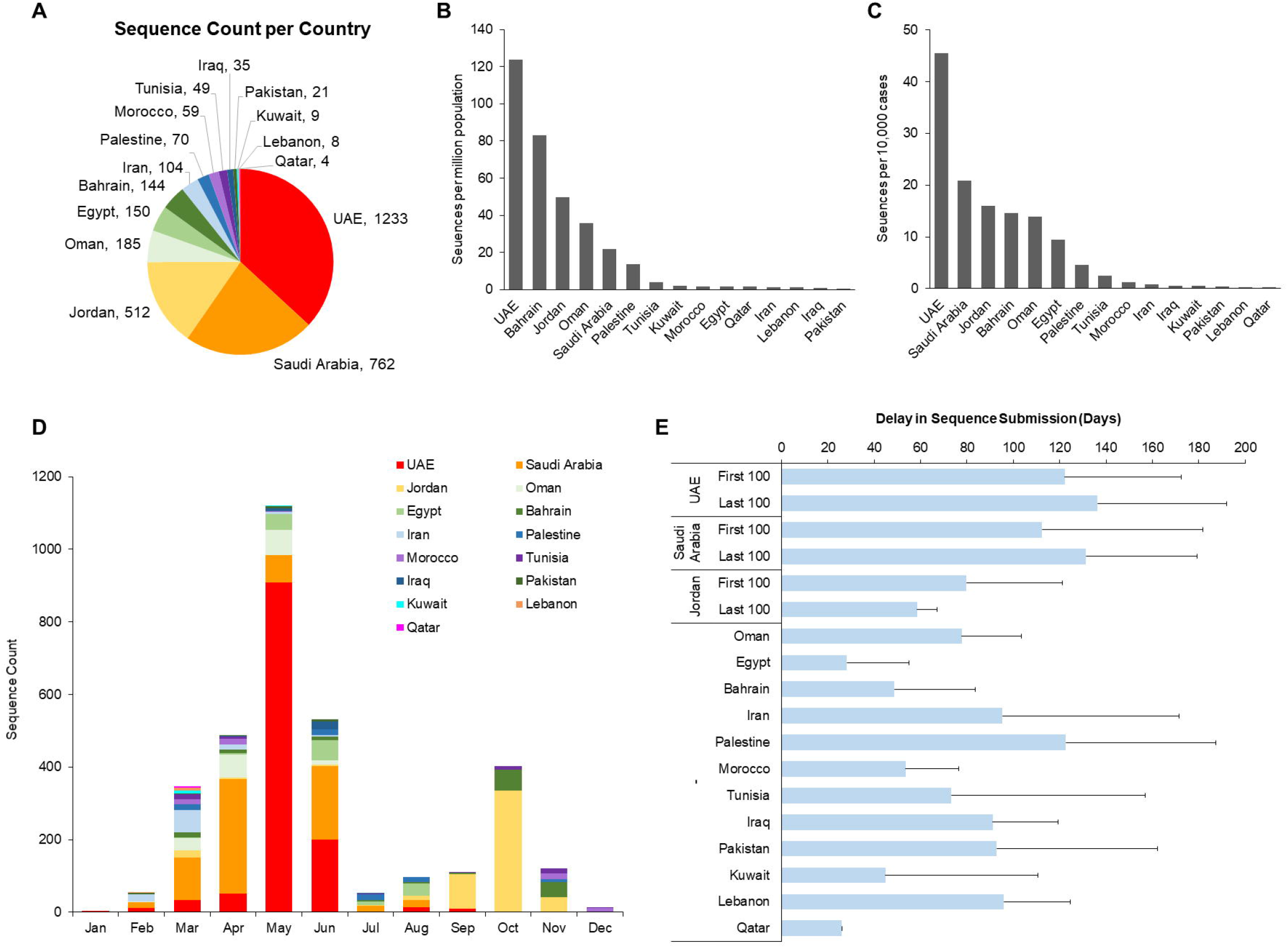
Distribution of SARS-CoV-2 sequences in EMR countries. (A) Pie chart showing number of available sequences as of 1 January 2021. (B) Histogram showing ratio of sequences per million population. (C) Histogram showing ratio of sequences per 10,000 confirmed cases of SARS-CoV-2. (D) Temporal distribution of sequence collection in EMR countries from January through December 2020. (E) Averages of delay in sequence submission (in days) of available EMR countries. For UAE, Saudi Arabia and Jordan, only the first and last 100 collected sequences were considered. Error bars represent standard deviation.

**Figure 1.**
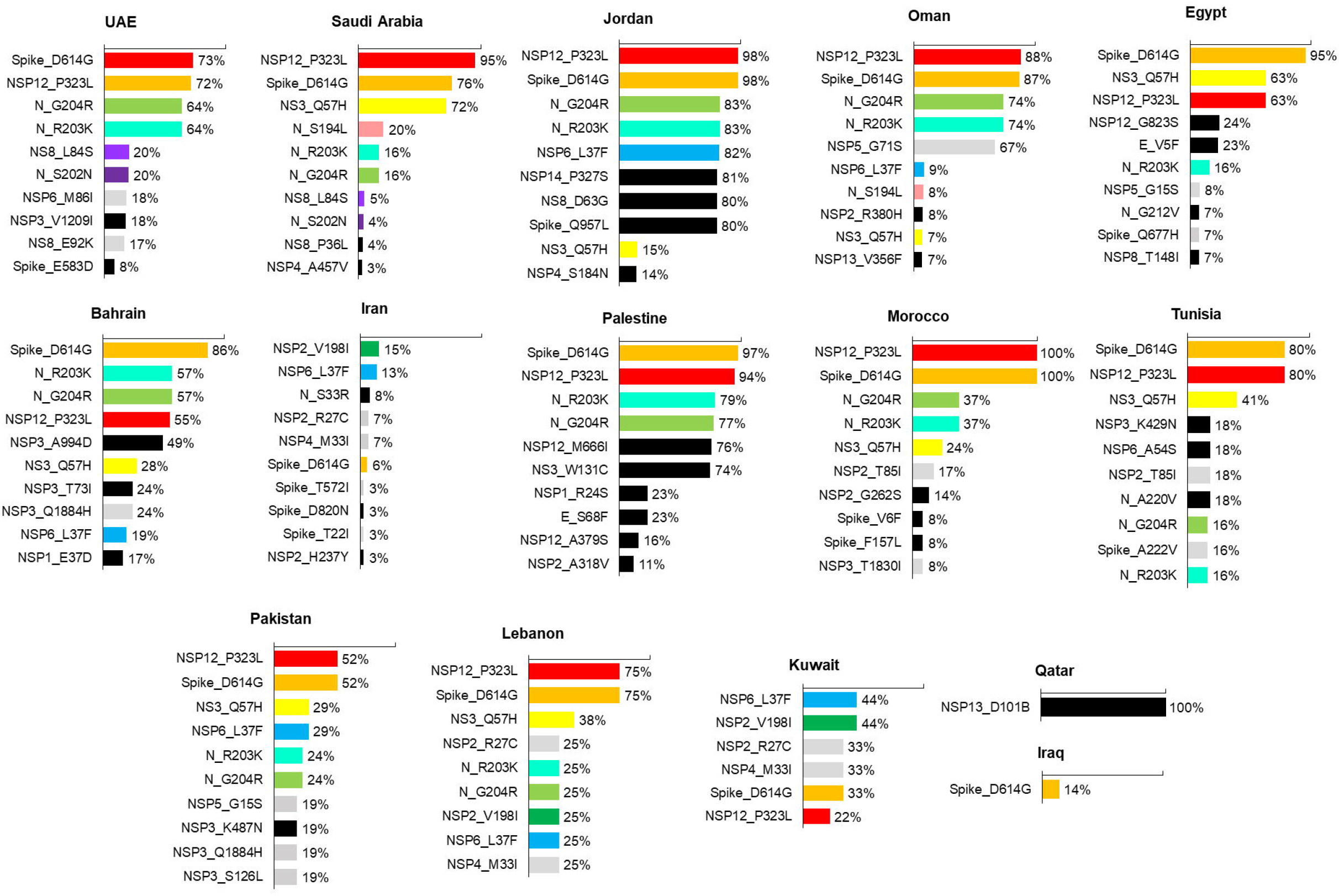
Common SARS-CoV-2 substitutions detected in EMR countries. Bar graphs showing percentages of most prevalent SARS-CoV-2 variants per country, excluding single-incidence variants. Colored are the top ten most represented substitutions; in black are country-specific ones.

## Results

We identified 3345 high-coverage SARS-CoV-2 genome sequences from EMR countries – no sequences were available for Afghanistan, Djibouti, Libya, Somalia, Sudan, Syria, and Yemen. Three countries accounted for ∼75% of the sequence count overall – UAE (n=1233), Saudi Arabia (n=762) and Jordan (n=512) (**Figure 1A**). All but 5 EMR countries – Iran, Tunisia, Iraq, Pakistan and Kuwait – had near-full length sequences of SARS-CoV-2 (**Supplementary Figure 1**). UAE had the highest sequence to population ratio (123.8 in 1 million), but also the highest number of sequences per reported COVID-19 cases (45.5 per 10,000 cases) (**Figure 1B, 1C**). When dates of collection were considered, none of the countries showed routine and consistent sequencing; most sequences were collected before July (e.g. from UAE, Saudi Arabia, Oman) while only few countries (mainly Jordan, Bahrain and Morocco) reported sequences from the last three months (**Figure 1D**). Moreover, a significant delay between sample collection and sequence submission was observed across all countries (ranging 30-150 days), with no noticeable progress between first and last 100 days for the top three represented countries (**Figure 1E)**.

Using CoVsurver from GISAID, amino acid substitutions were identified in each viral protein sequence and the top ten prevalent variants for each country were then plotted (excluding single-incidence variants) (**Figure 2**). In total, 2155 non-synonymous mutations were detected in EMR countries, out of which ten common variants were found to be relatively most common (i.e. in ≥ 8 EMR countries and > 2.5% of all sequences). These are: NSP12_P323L, S_D614G, NS3_Q57H, N_R203K, N_G204R, N_S194L, NSP6_L37F, NSP2_V198L, NS8_L84S and N_S202N. Besides, many country-specific substitutions were detected (here defined as found in 3 other countries or less, and not listed in other countries’ most common mutations). For instance, NSP3_V1209I was found in 18% of UAE’s sequences, Spike_Q957L in 80% of Jordan’s sequences and NSP12_M666I in 76% of Palestine’s sequences (**Figure 2**). NSP12_P323L and S_D614G were predominantly detected in 10 out of 15 countries. S_D614G was also dominant in Bahrain. In Kuwait and Iran, NSP6_L37F and NSP2_V198L were the most frequently reported variants.

**Figure 2.**
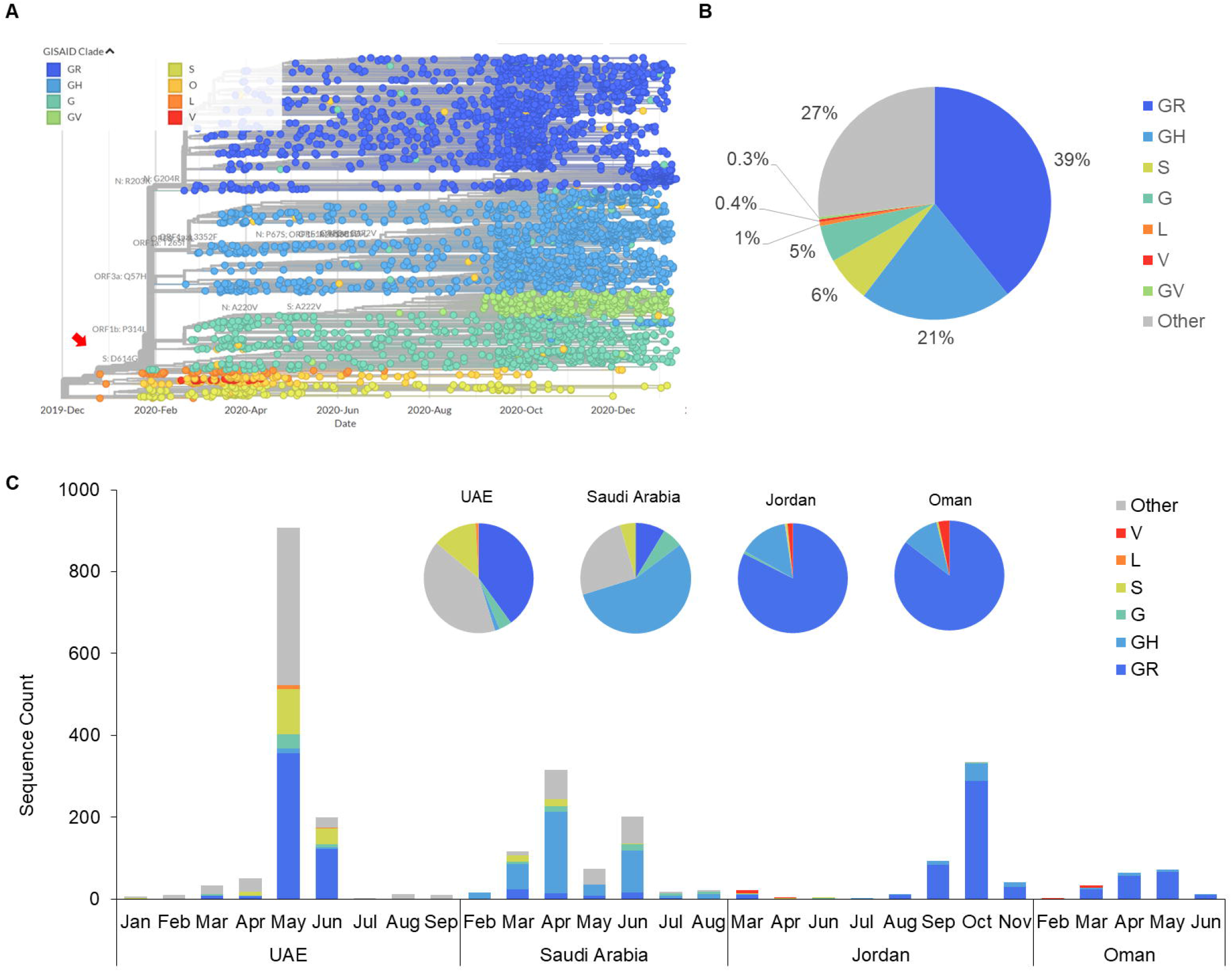
EMR sequences are dominantly distributed in G, GR and GH GISAID clades. (A) Phylogenetic tree of GISAID clades plotted over time. Red arrow points to the introduction of S_D614G, marking the emergence of G, GR and GH clades (reproduced from Nextstrain [Ref. 17]. (B) Clade distribution of all 3345 sequences. (C) Temporal distribution of sequences as clades in the top four represented countries.

Next, we used the Nextclade tool from Nextstrain to assign clades (GISAID nomenclature) to the sequences (**Figure 3A)** (GISAID, 2020). Of all sequences from EMR countries, 65.8% belonged to the G (20A per Nextstrain clades), GR (20B), GH (20C) and GV (20A.EU1) clades, which emerged following the introduction of the S_D614G variant (**Figure 3B**, (Alm et al., 2020)).

Moreover, these clades were almost exclusively represented as early as February and March in Saudi Arabia. In UAE, Jordan and Oman, clade distribution was heterogeneous with multiple clades cocirculating between February through November (**Figure 3C**).

For the sequence cohort submitted by 12 November 2020, more specimens were sequenced from males in the EMR (2.05:1 male-to-female in top four represented countries, independent t-test p < 0.004): Saudi Arabia (4.44:1), UAE (2.25:1), Oman (1.42:1) and Egypt (1.49:1) (**Figure 4A**). No statistically significant difference in gender distribution of the top represented SARS-CoV-2 variants were observed in these countries (independent t-test for all variants, p > 0.270) (**Figure 4B**). By analyzing the age distribution of the top variants in these countries, we observed a statistically significant underrepresentation in the number of sequenced viruses from children (0-9 years) (3.43% ± 1.21 S.E.M.) and young adolescents (10-18 years) (2.26% ± 1.16), while adults (19-49) were more represented (62.5% ± 8.84) (ANOVA; F = 73.4, *p* = 1.15E-9) (**Figure 4C**). For the three remaining age categories (19-49, 50-59 and >60 years), we report no statistically significant difference in the occurrence of the Wuhan-like or any of the identified variants (multivariate ANOVA for all variants, F < 0.906, p > 0.487) (**Figure 4D**).

**Figure 4.**
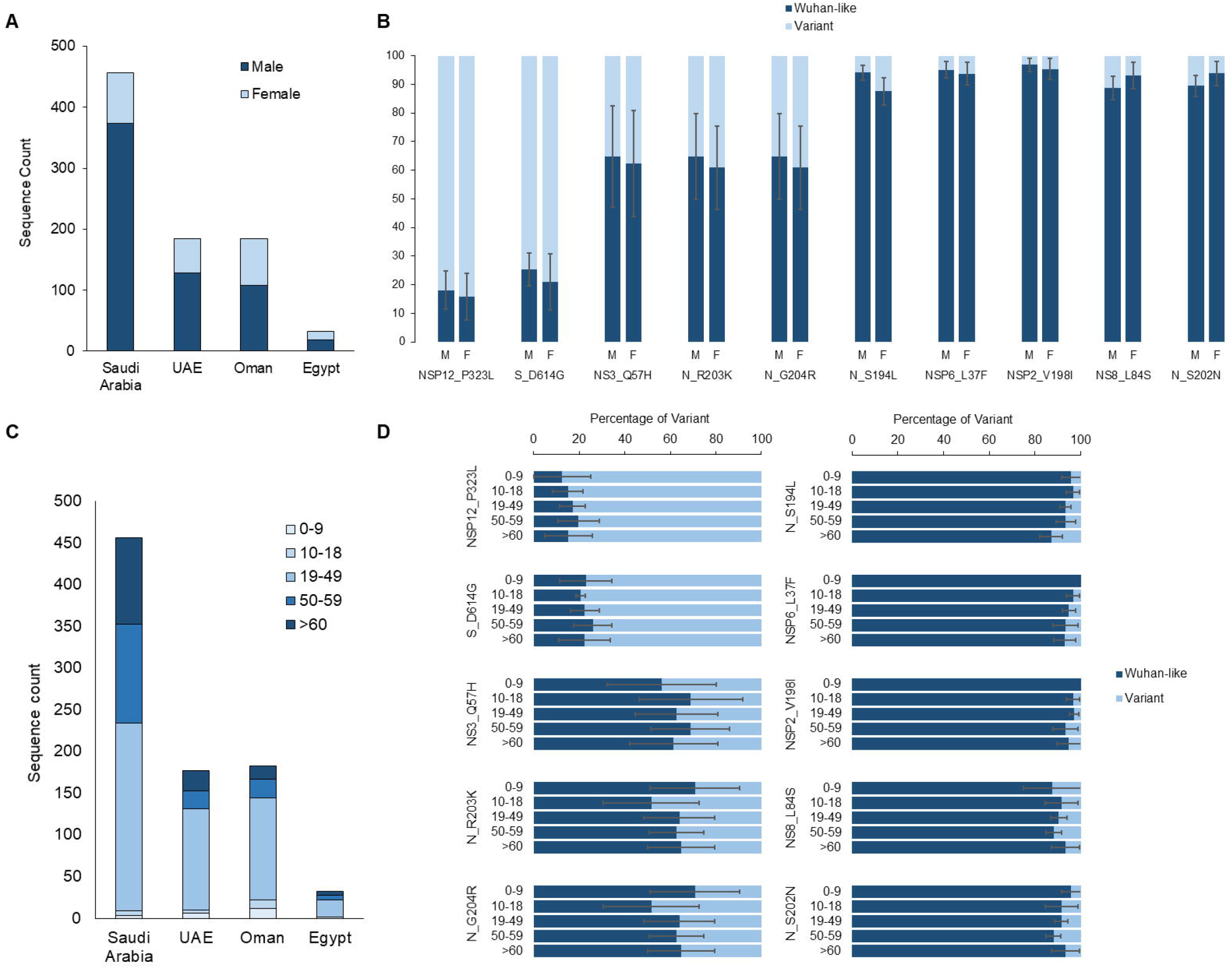
Gender and age distribution of SARS-CoV-2 variants in four EMR countries. Gender distribution of (A) sequences of Saudi Arabia, UAE, Oman and Egypt and (B) select top most frequent variants of these countries. (C) Age groups distribution of sequences from four EMR countries and (D) of most frequent variants. Sequences in this figure are as of 12 November 2020 due to the inability to extract demographics from metafiles after this date.

We next sought to determine whether a correlation exists between the incidence of the ten most common variants and countries’ CFRs. Only seven countries were included in this analysis – UAE, Saudi Arabia, Jordan, Oman, Bahrain, Palestine and Morocco – where only NSP12_P323L out of the ten variants was significantly correlated with an increase in CFR (R=0.883, *P=*0.008) (**Figure 5**). However, consistent with previous studies (Ilmjärv et al., 2020; Kannan et al., 2020), we confirm that NSP12_P323L seems to have quickly co-evolved following S_D614G introduction, as they are more often co-detected among SARS-CoV-2 genome sequences (**Figure 6A**). This double variant ‘D614G & P323L’ is found to be evenly represented among different age groups in UAE, Saudi Arabia, Oman and Egypt (for sequences submitted by 12 November) (**Figure 6B**). After repeating the correlation analysis while accounting for the single versus double occurrence of D614G and P323L, we found that neither single nor double variants were significantly correlated with CFR. The P323L-single variant was mainly detected in Saudi Arabia (19% of it sequences) and among 0.5% sequences reported from UAE, while in the other countries it was exclusively co-detected with D614G. As such, we suggest that the earlier correlation of NSP12 P323L is likely the result of considering single- and double-variants as one group, specifically in Saudi Arabia. However, when such distinction was made, no correlation with higher CFR was reported in either group (**Figure 6C**).

**Figure 5.**
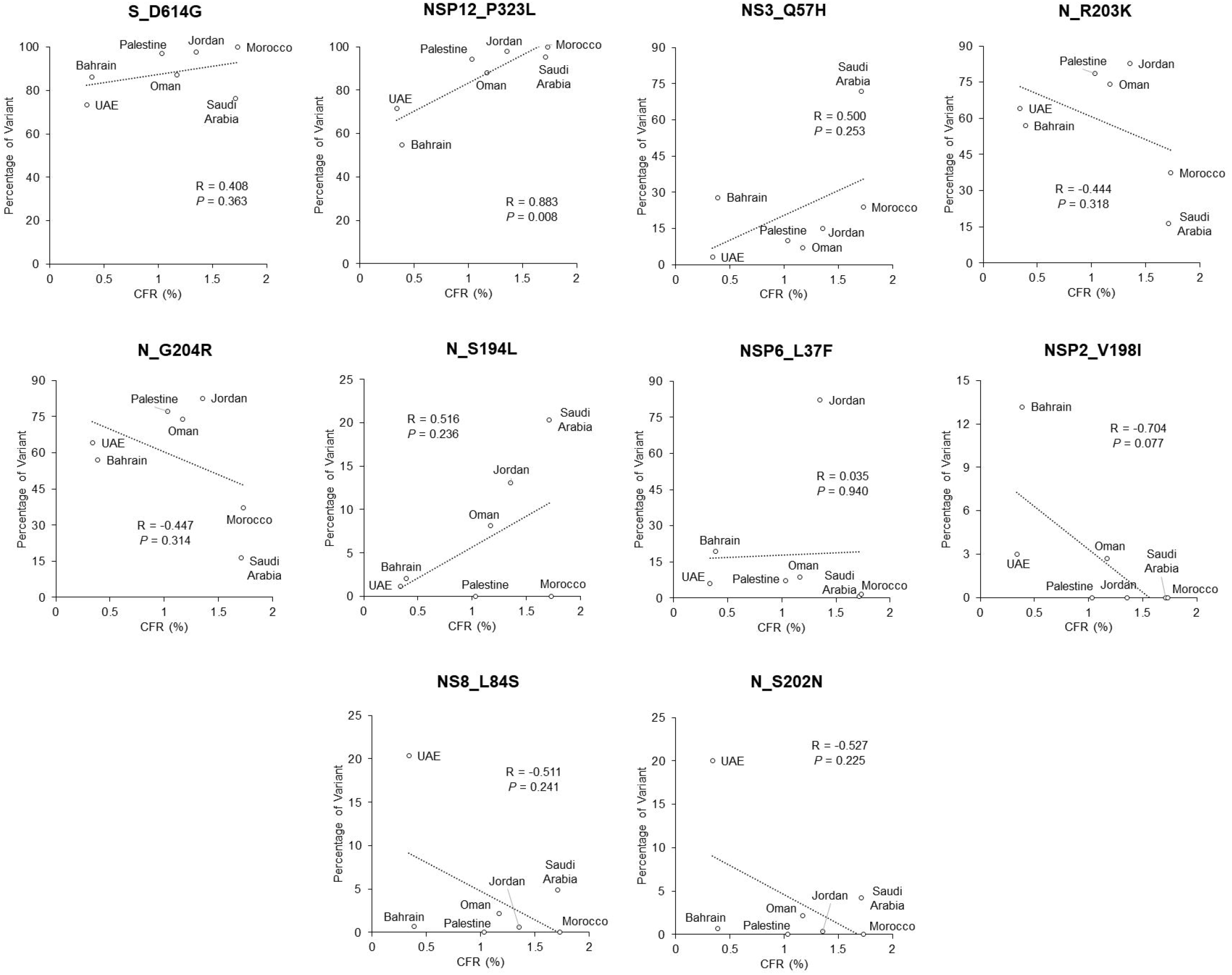
Only NSP12_P323L of the ten most common SARS-CoV-2 single variants correlates with higher case fatality rates in seven EMR countries. Scatter plots showing percentages of each of the top ten represented substitutions with cumulative CFR as of 1 January 2021. R, Pearson’s correlation coefficient.

**Figure 6.**
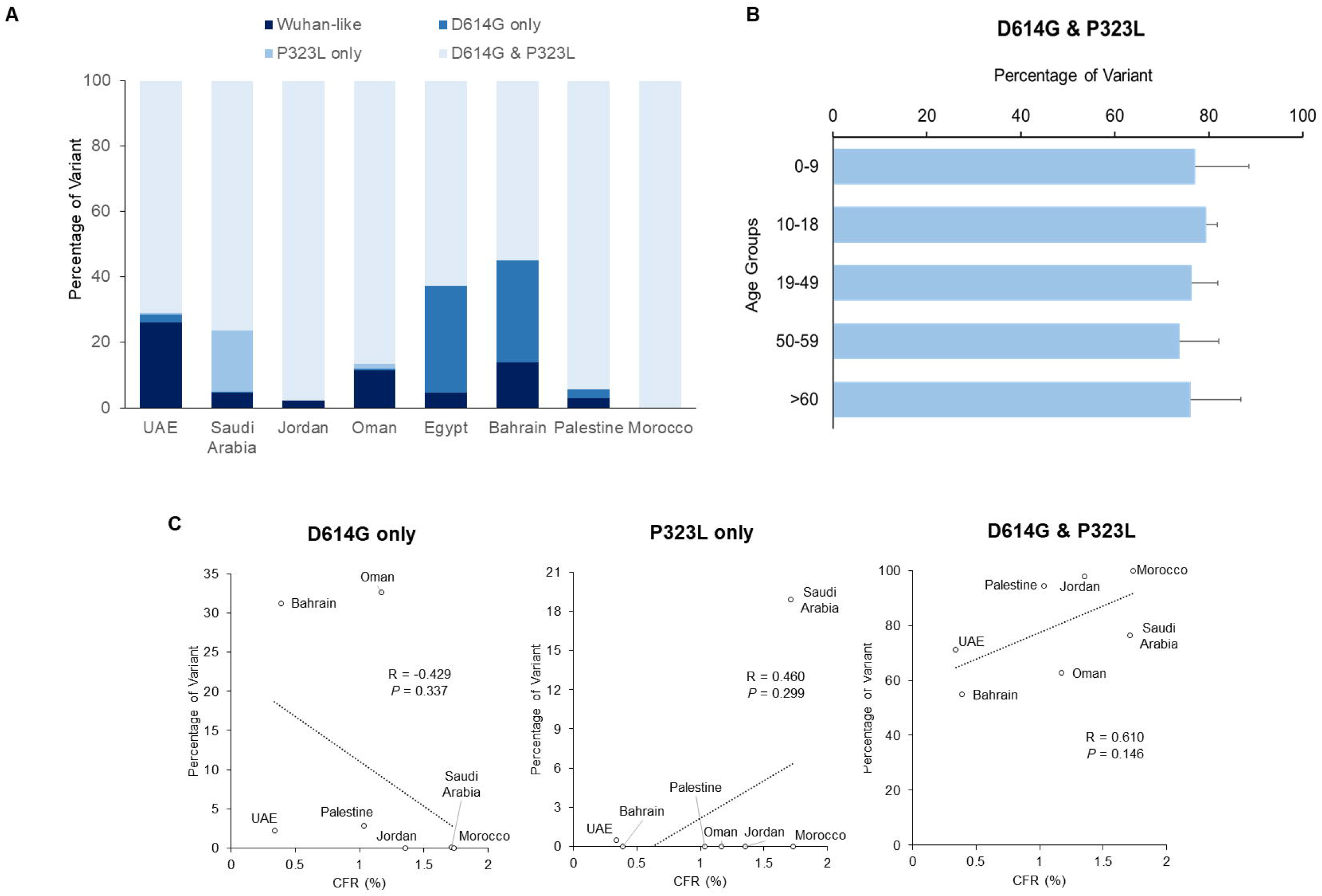
The ‘D614G & P323L’ double variant is predominant in select EMR countries, but does not correlate with increased fatality. (A) Percentages of Wuhan-like, S_D614G only, NSP_P323L only and ‘D614G & P323L’ double variants in eight EMR countries. (B) Distribution of the ‘D614 & P323L’ variant based on age groups in top four represented EMR countries. (C) Scatter plots showing correlation of three variants with CFR in seven EMR countries. R, Pearson’s correlation coefficient.

Since sequences of UAE, Saudi Arabia and Oman were dominantly collected before September and those of Jordan and Bahrain were collected afterwards (**Figure 1D**), we repeated our analysis but this time accounting for the cumulative incidence of the ‘D614G & P323L’ double variant and the CFR at the end of each month. We observed no significant association between these two variables from March through September (**Supplementary Figure 2**). To further investigate this correlation, we considered the cumulative monthly incidence of ‘D614G & P323L’ and the monthly CFRs (**Figure 7A**): while UAE showed a significantly negative correlation between ‘D614G & P323L’ incidence and the monthly CFR (R=-0.830, *P=*0.021), Saudi Arabia showed a significantly positive correlation (R=0.909, *P*=0.012). Meanwhile, Jordan and Oman had no significant associations (R=-0.101, *P=*0.849; R=0.921, *P*=0.079 respectively) (**Figure 7B**). This suggests that by accounting for the change in CFR for each country with time the correlation between ‘D614G & P323L’ incidence and higher fatality does not necessarily hold true contrary to previous findings (Becerra-Flores and Cardozo, 2020; Eaaswarkhanth et al., 2020; Toyoshima et al., 2020).

**Figure 7.**
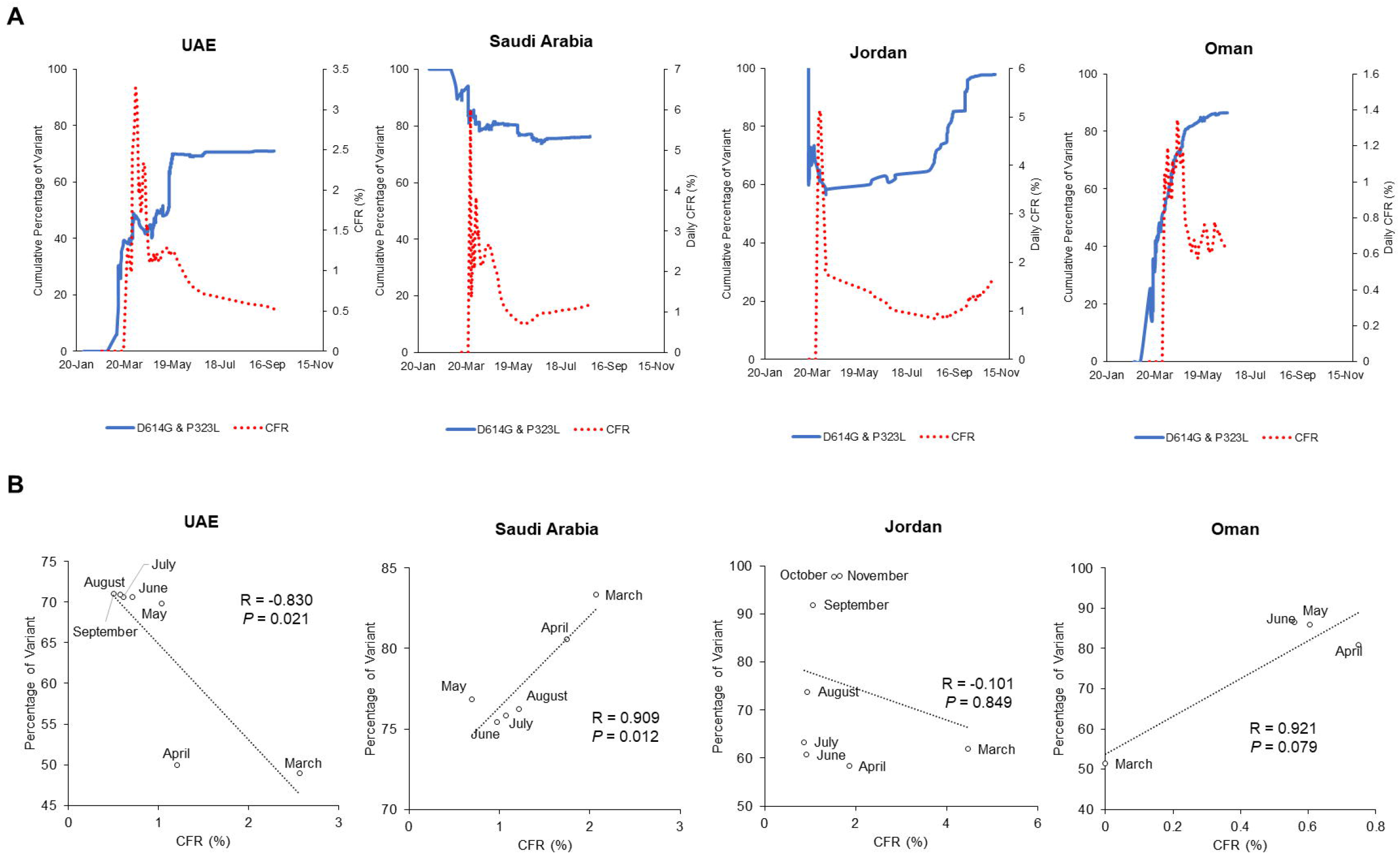
Temporal distribution and fatality correlation of the ‘D614G & P323L’ double variant with CFR in each of the top four represented EMR Countries. (A) Time-based evolution of double variant percentages and CFR in Saudi Arabia, UAE, Oman and Egypt. (B) Scatter plots showing correlations of the ‘D614G & P323L’ double variant with CFR over select months of pandemic life in the top four represented EMR countries. R, Pearson’s correlation coefficient.

## Discussion

In this study, we analyzed whole-genome sequences of SARS-CoV-2 collected in the EMR countries. Significant data gaps are notable, whereby sequences were lacking for seven EMR countries – the majority of which are in war conflict zones (e.g., Syria, Libya, and Yemen (Daw, 2020)). In general, large disparities in the availability of sequences are observed across EMR. Only five countries reported ≥150 sequences spanning the first year of the pandemic, with UAE, Saudi Arabia and Jordan exceeding 500 sequence submissions. These data gaps highlight the challenges for monitoring the pandemic in developing countries particularly in conflict zones. It also emphasizes the importance of building regional sequencing capacities for timely and continuous monitoring of viral evolution during the pandemic (Al Saidi et al., 2020; Salameh, 2020).

Despite the diversity in the substitutions detected across countries, only a handful of variants predominated. For instance, a mere 9 out of 945 non-synonymous mutations (0.95%) in UAE and 6 out of 421 mutations (1.4%) in Saudi Arabia were detected in more than 10% of the respective total of sequences (**Figure 2**). This suggests that consistent with other studies, most SARS-CoV-2 mutations are neutral or deleterious (van Dorp et al., 2020a). Although a random founder’s effect cannot be ruled out, the increased frequency of certain substitutions might be attributable to a fitness advantage (Callaway, 2020; Grubaugh et al., 2020; Kannan et al., 2020; Plante et al., 2020; Yurkovetskiy et al., 2020; Zhang et al., 2020). For instance, the S_D614G was found to predominate all viral strains globally (78% of sequences by May 2020) and was associated with increased viral loads in the upper respiratory tract of COVID-19 patients (Korber et al., 2020). Along the same line, Plante et al. recently demonstrated that S_D614G enhanced viral replication and infectivity of SARS-CoV-2 in human lung epithelial cells and primary human airway tissues and increased infectious virus loads in hamsters’ nasal cavities (Plante et al., 2020). Hou et al. further showed that the S_D614G variant displayed faster transmission and increased competitive fitness *in vivo* compared to wild-type virus in hamsters (Hou et al., 2020). These findings were also confirmed in hACE2 knock-in mice as well as in ferrets (Zhou et al., 2021). On a molecular level, S_D614G allows for more efficient infectivity by shifting the spike protein to a more ‘open’ conformation that is more favorable of ACE2 binding (Yurkovetskiy et al., 2020). Interestingly, however, a recent analysis by van Dorp et al. could not identify any single recurrent mutation, including the S_D614G, to be convincingly associated with increased viral transmission in humans (van Dorp et al., 2020b).

Several studies reported a positive correlation between S_D614G incidence and mortality (CFR) (Becerra-Flores and Cardozo, 2020; Eaaswarkhanth et al., 2020; Toyoshima et al., 2020). However, a more recent study of 25,000 SARS-CoV-2 sequences in the United Kingdom did not associate the incidence of this mutation with higher COVID-19 mortality or with clinical severity in patients (Volz et al., 2021). Similarly, Korber et al. did no find an association between S_D614G and increased disease severity or hospitalization (Korber et al., 2020). In our sequence set, we saw no association between S_D614G with CFRs of select EMR countries. Instead, the increasing incidence of NSP12_P323L (of the ten most common substiutions), when considered alone, was correlated with a higher mortality rate. However, our data as well as others’ (Ilmjärv et al., 2020; Kannan et al., 2020) show that the NSP12_P323L and S_D614G are frequently coupled in most sequences, suggesting co-evolution of these variants (**Figure 6A**). Therefore, we assessed the combined effect of the ‘D614G & P323L’ double variant on mortality. We found no correlation between the cumulative incidence of the ‘D614G & P323L’ variant and CFRs in the EMR countries, consistent with earlier observations that found no association between ‘D614G & P323L’ and deaths per million, CFR and patient status (Ilmjärv et al., 2020). In contrast to the well-studied S_D614G, the effect of NSP12_P323L on viral fitness is much less understood (Hillen et al., 2020; Pachetti et al., 2020). One study speculated that it negatively impacts NSP8’s association with RdRp leading to an attenuated polymerase activity (Ilmjärv et al., 2020), while another suggested that it enhances hydrophobic NSP8-RdRp interactions leading to improved processivity of RdRp and enhanced viral replication overall (Kannan et al., 2020). In all cases, it is speculated that the ‘epidemiological success’ of the G, GH, GR and GV lineages could be attributed to the combined viral fitness advantage conferred by both NSP12_P323L and S_D614G (Ilmjärv et al., 2020).

Considering the limitations due to the discrepant number of sequences available (as mentioned above) or to country-specific variations in managing the COVID-19 pandemic (such as testing capacity, age stratification, enforcing/relaxing lockdown policies, etc.), we assessed the association of ‘D614G & P323L’ incidence with CFR within each of the top four represented countries over time. Interestingly, we found a significantly negative correlation between ‘D614G & P323L’ incidence and COVID-19 CFR in UAE, but an otherwise positive correlation in Saudi Arabia (**Figure 7B**). This suggests that the reported correlation between these variants and increased CFR might be a mere coincidence rather than true causation. Noteworthy, our method of calculating daily/monthly CFR is credited for accounting for a 13-day lag between cumulative cases and deaths (Wilson et al., 2020). Yet, it overestimates the case fatality risk early on in the pandemic (Ge and Sun, 2020), a limitation that would apply to our final analysis. Then again, considering the overwhelming discrepancy in fatality correlations between countries, it is unlikely that early bias in CFR estimation would have altered our conclusion. Additionally, a more accurate method to estimate COVID-19 mortality would be the infection fatality rate (IFR), inferred from seroprevalence studies. However, such data is only available for Iran, Pakistan, and Qatar out of all EMR countries according to a recent WHO review (Ioannidis, 2020b).

Other genomic mutations were associated with milder disease, such as a major deletion (Δ382; nucleotide positions: 27,848 to 28,229), which results in truncated ORF7b and arrested transcription of ORF8 (Su et al., 2020). Patients infected with the SARS-CoV-2 Δ382 variant had less systemic release of proinflammatory cytokines and better clinical outcomes (i.e. lower frequency of hypoxia requiring supplemental oxygen) than wild-type (Young et al., 2020). This variant, however, was not present in any of our sequence cohort. Moreover, an S_ N439K variant was reported to enhance spike binding to its protein hACE2, resulting in immune escape from a panel of neutralizing monoclonal antibodies (Thomson et al., 2021). We detected only one sequence carrying this mutation from Morocco (EPI_ISL_728353).

On a final note, the COVID-19 pandemic cannot be viewed in the absence of other respiratory viruses, especially in view of the increased concerns regarding a ‘Twindemic’ (Uyeki et al., 2020; Adlhoch et al., 2021). In the case of influenza, few cases were reported in EMR countries in 2020-21 season (<20 positive specimens per week; GISRS report), a pattern that was established worldwide (Jones, 2020). Previous studies from the United States and Australia reported a co-infection rate ranging from 2% to 6.9% between SARS-CoV-2 and other respiratory viruses including rhinovirus/enterovirus, RSV, other coronaviruses, and influenza virus (Kim et al., 2020; Kim et al., 2021). Elghoudi et al. reported an incidence of 3.1% (9/288) respiratory co-infections with SARS-CoV-2 in children and young adolescents in a hospital in UAE (Elghoudi et al., 2020). In addition to co-infections with respiratory viruses a study from Pakistan reported a relatively high incidence of co-infections between SARS-CoV-2 and the endemic dengue virus – 25% (5/20) of studied samples (Saddique et al., 2020). A meta-analysis study estimated that while 3% of COVID-19 patients (out of 1014 patients from 16 studies) had a confirmed viral co-infection, there was no significant increase in hospitalization or ICU admission in the co-infected cohort (Lansbury et al., 2020). Overall, the incidence of respiratory viral co-infections is low and may not be considered a key factor associated with mortality.

Our analysis reveals the diversity of SARS-CoV-2 in EMR and emphasizes the importance of considering temporal incidence and concurrent mutations in disease correlations. Our study is limited by the relatively low number of specimens sequenced per country spanning ten months of COVID-19 pandemic. There is a critical need for continuous and consistent surveillance of SARS-CoV-2 genomic variations in the EMR region.

## Supporting information

GISAID Acknowledgement

## Data Availability

SARS-CoV-2 sequences were downloaded from GISAID EpiCoV database as of 1 January 2021 for EMR countries.

## Acknowledgments

S.O. is the recipient of a joint doctoral fellowship from the American University of Beirut (AUB) and the National Council for Scientific Research of Lebanon (CNRS-L). We gratefully acknowledge the authors from the originating laboratories responsible for obtaining the specimens, as well as the submitting laboratories where the genome data were generated and shared via GISAID, on which this research is based (a detailed list of authors and laboratories can be found in the supplementary material). We also thank Dr. Colin A. Smith and Dr. Heinrich zu Dohnna (Biology department, American University of Beirut) for their valuable comments.

## Competing Interests

The authors declare no conflicts of interest.

## Funding

This research did not receive any specific grant from funding agencies in the public, commercial, or not-for-profit sectors.

## Figure Legends

**Supplementary Figure 1. Genome length of available EMR sequences**. Sequence size distribution (%) of available EMR sequences in two categories: <5000 bp and >25000 bp.

**Supplementary Figure 2. Correlation between the ‘D614G & P323L’ double variant with CFR over eight months of the pandemic**. Scatter plots showing cumulative percentages of the double variant in seven EMR countries versus respective CFR as of the end of each month, March through September and November. R, Pearson’s correlation coefficient.

## Notes

### Competing Interest Statement

The authors have declared no competing interest.

### Summary of Updates

We updated EMR sequences as of 1 Jan 2021.

