## Supplementary figures and images for "No association between the SARS-CoV-2 variants and mortality rates in the Eastern Mediterranean Region"

### Supplementary Figure 1

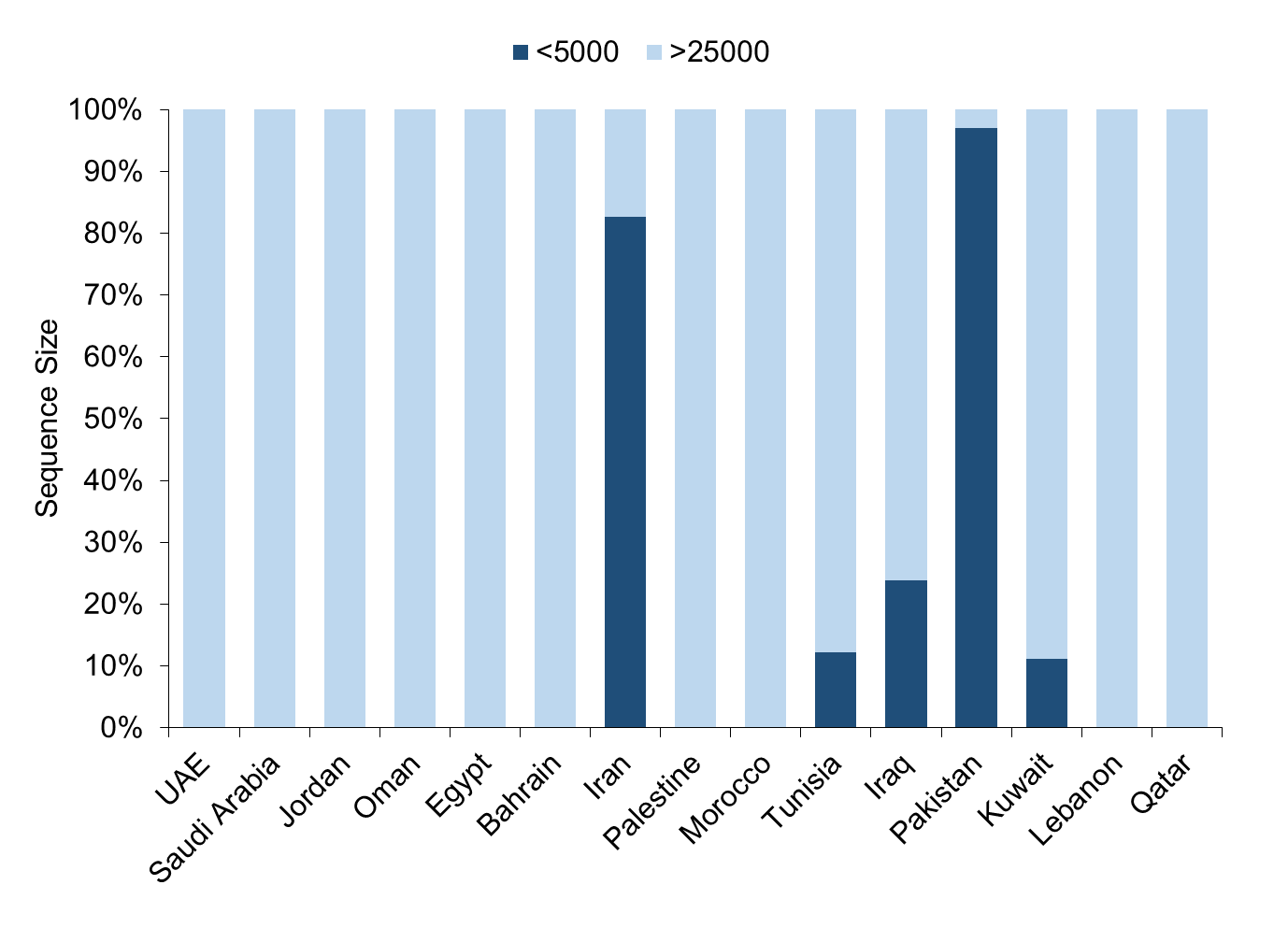

### Supplementary Figure 2

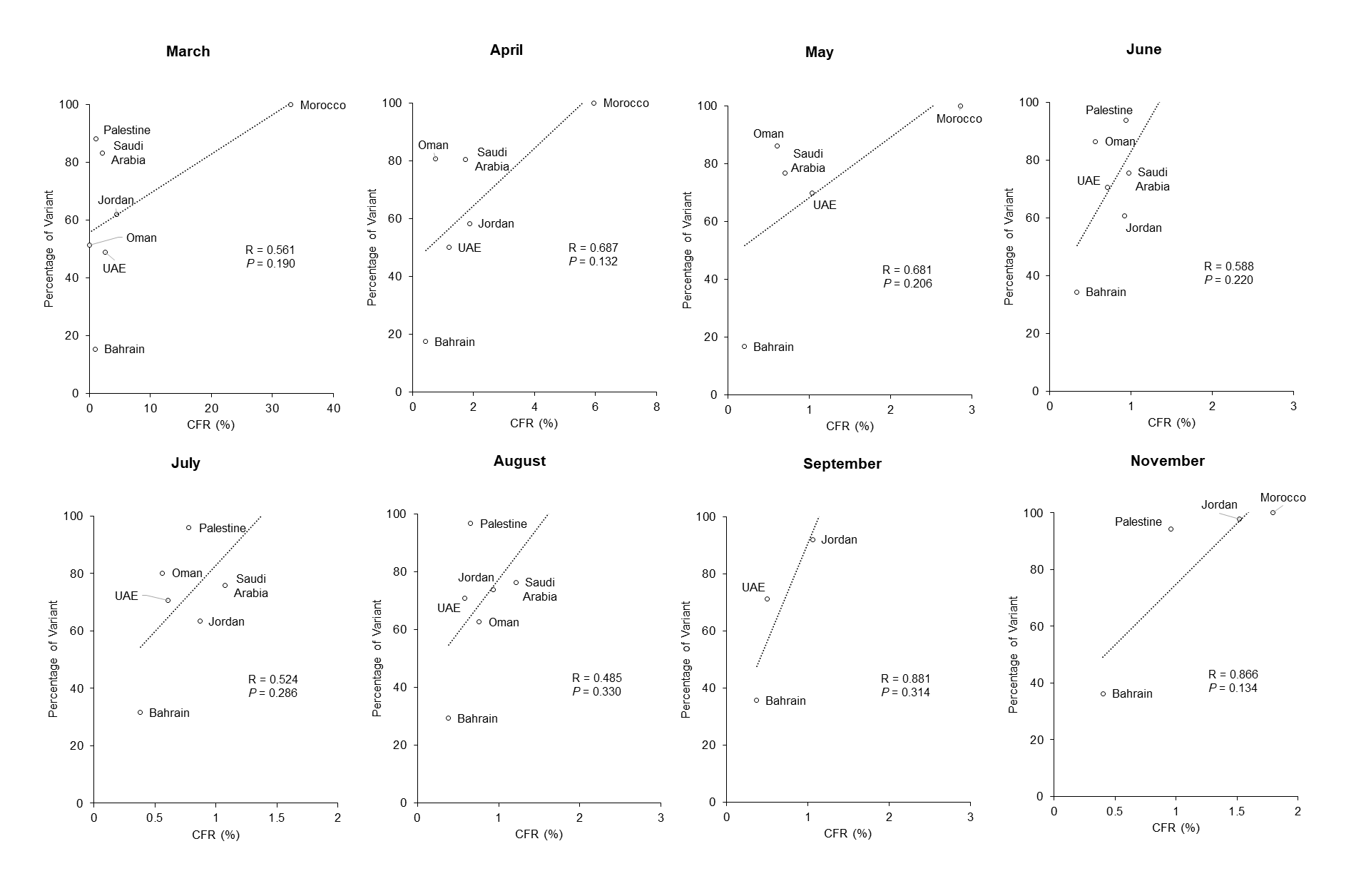
